# What is the effectiveness of financial support schemes for individuals requested to self-isolate following a positive Covid test or positive contact: A rapid review

**DOI:** 10.1101/2022.09.15.22279969

**Authors:** Tom Winfield, Lauren Elston, Jenni Washington, Elise Hasler, Ruth Lewis, Alison Cooper, Adrian Edwards

## Abstract

Testing for COVID-19 has been deployed globally as a tool to interrupt transmission through isolating positive contacts from the broader population. Financial support systems have been deployed to increase the isolation compliance, there is uncertainty as to the effectiveness of these measures.

Three reviews were identified, as well as four primary studies that were published after the review search dates.

Six studies showed that financial support for isolation was associated with a higher compliance to isolate. Two epidemiological modelling studies found that increased levels of social isolation were associated with a reduction in COVID-19 transmission. The findings from a DCE demonstrated a positive relationship with longer isolation duration and higher financial requirements. An economic model showed that support programmes have the potential to be a cost-effective intervention. A retrospective observational study offered evidence supporting the viability of delivering medically assisted isolation hotels for people unable to isolate at home. Further to the COVID-19 literature, two household surveys found that financial support and improved social restriction information was associated with compliance with H1N1 isolation

Policy and practice implications: There is limited evidence to suggest that financial support for isolation can increase compliance, lower social engagement, and reduce infection levels. There is insufficient evidence to inform the optimal scale of financial support required. There was no evidence related to effectiveness of financial support for disadvantaged populations who are required to isolate or any insight to the impact of financial support on equality

The overall certainty in the evidence is relatively low. Most studies relied on participant reported data on preference or behaviour, and where observational data were used there were issues with data quality and unobserved cofounders.

Rapid Review Details

Review conducted by
Health Technology Wales

Review Team
Lauren Elston, Jenni Washington, Elise Hasler, Tom Winfield

Review submitted to the WCEC on
27^th^ July 2022

Stakeholder consultation meeting
13^th^ June 2022

Rapid Review report issued by the WCEC on
August 2022

WCEC Team

- Adrian Edwards, Alison Cooper, Ruth Lewis, Jane Greenwell and Micaela Gal involved in drafting Topline Summary and editing

This review should be cited as
RR00020.Wales COVID-19 Evidence Centre. A rapid review of the effectiveness of financial support schemes for individuals requested to self-isolate following a positive Covid test or positive contact. August 2022
This report can be accessed from the WCEC library: https://healthandcareresearchwales.org/wales-covid-19-evidence-centre-report-library

Disclaimer
The views expressed in this publication are those of the authors, not necessarily Health and Care Research Wales. The WCEC and authors of this work declare that they have no conflict of interest.

TOPLINE SUMMARY

What is a Rapid Review?
Our rapid reviews use a variation of the systematic review approach, abbreviating or omitting some components to generate the evidence to inform stakeholders promptly whilst maintaining attention to bias. They follow the methodological recommendations and minimum standards for conducting and reporting rapid reviews, including a structured protocol, systematic search, screening, data extraction, critical appraisal, and evidence synthesis to answer a specific question and identify key research gaps. They take 1-2 months, depending on the breadth and complexity of the research topic/ question(s), extent of the evidence base, and type of analysis required for synthesis.

Who is this summary for?
Welsh Government

Background / Aim of Rapid Review
Testing for COVID-19 has been deployed globally as a tool to interrupt transmission through isolating positive contacts from the broader population. Financial support systems have been deployed to increase the isolation compliance, there is uncertainty as to the effectiveness of these measures.

Key Findings
Three reviews were identified, as well as four primary studies that were published after the review search dates. Due to the diversity and paucity of evidence identified, the primary studies included in the reviews (n = 5) were extracted and reported alongside the other primary evidence. This resulted in 9 primary studies extracted and summarised in this report.

Extent of the evidence base

- The primary studies focused mainly on the COVID 19 pandemic (n=7) with two studies set in the context of the H1N1 pandemic.
- The study types included: epidemiological modelling studies (n=2), economic modelling study (n=1), questionnaire-based publication (n=1), discrete choice experiments (DCEs) (n=2), retrospective observational study (n=1), and household surveys (both H1N1, n=2).
- The studies were conducted in the USA (n=3), Brazil (n=1), Iran (n=1), Australia (n=2, H1N1 studies), or across multiple countries (USA, Mexico, and Kenya; n=1). No UK-based studies were identified.
- Most studies (n=7) included a general population, but one study focused on a homeless population, and one study included staff and students at university.

Recency of the evidence base

- 7 primary studies were conducted in the last 2 years; the 2 studies from the H1N1 pandemic were conducted in 2011-12.

Evidence of effectiveness

- Six studies showed that financial support for isolation was associated with a higher compliance to isolate.
- Two epidemiological modelling studies found that increased levels of social isolation were associated with a reduction in COVID-19 transmission.
- The findings from a DCE demonstrated a positive relationship with longer isolation duration and higher financial requirements.
- An economic model showed that support programmes have the potential to be a cost-effective intervention.
- A retrospective observational study offered evidence supporting the viability of delivering medically assisted isolation hotels for people unable to isolate at home.
- Further to the COVID-19 literature, two household surveys found that financial support and improved social restriction information was associated with compliance with H1N1 isolation.

Policy Implications

- There is limited evidence to suggest that financial support for isolation can increase compliance, lower social engagement, and reduce infection levels.
- There is insufficient evidence to inform the optimal scale of financial support required.
- There was no evidence related to effectiveness of financial support for disadvantaged populations who are required to isolate or any insight to the impact of financial support on equality

Strength of Evidence
The overall certainty in the evidence is relatively low. Most studies relied on participant reported data on preference or behaviour, and where observational data were used there were issues with data quality and unobserved cofounders.

## 1. BACKGROUND

This Rapid Review was conducted as part of the Wales COVID-19 Evidence Centre Work Programme. The above question was developed through collaboration with a range of stakeholders including from the Welsh Government, the WCEC Core Team, and Health Technology Wales.

### 1.1 Purpose of this review

Testing for COVID-19 has been deployed globally as a tool to interrupt transmission through isolating positive contacts from the broader population. Financial support systems have been deployed to increase isolation compliance, there is uncertainty as to the effectiveness of these measures.

## 2. RESULTS

### 2.1 Overview of the Evidence Base

Preliminary scoping searches identified three reviews that were relevant to this review question: two rapid reviews (Cardwell et al. 2022, Patel et al. 2021) and one systematic review (Bahji et al. 2021). A summary of the included reviews is provided in Table 1. Cardwell et al. (2022) identified and assessed the effectiveness of measures to support people in isolation or quarantine during the COVID-19 pandemic, and reported on two primary studies (Kavanagh et al. 2011, Kavanagh et al. 2012). Patel et al. (2021) evaluated the effectiveness of financial support interventions and reported on two studies (Bodas & Peleg 2020, Pichler et al. 2020). The third review by Bahji et al. (2021) aimed to evaluate strategies to aid self-isolation and quarantine for individuals with severe and persistent mental illness during the COVID-19 pandemic, and included one relevant study (Fuchs et al. 2021). All five studies identified by these reviews were unique, and no studies overlapped between the reviews. Due to the lack of primary evidence identified for this report, each of these five studies have been extracted and reported individually alongside the other primary evidence in Table 2.

**Table 1:**
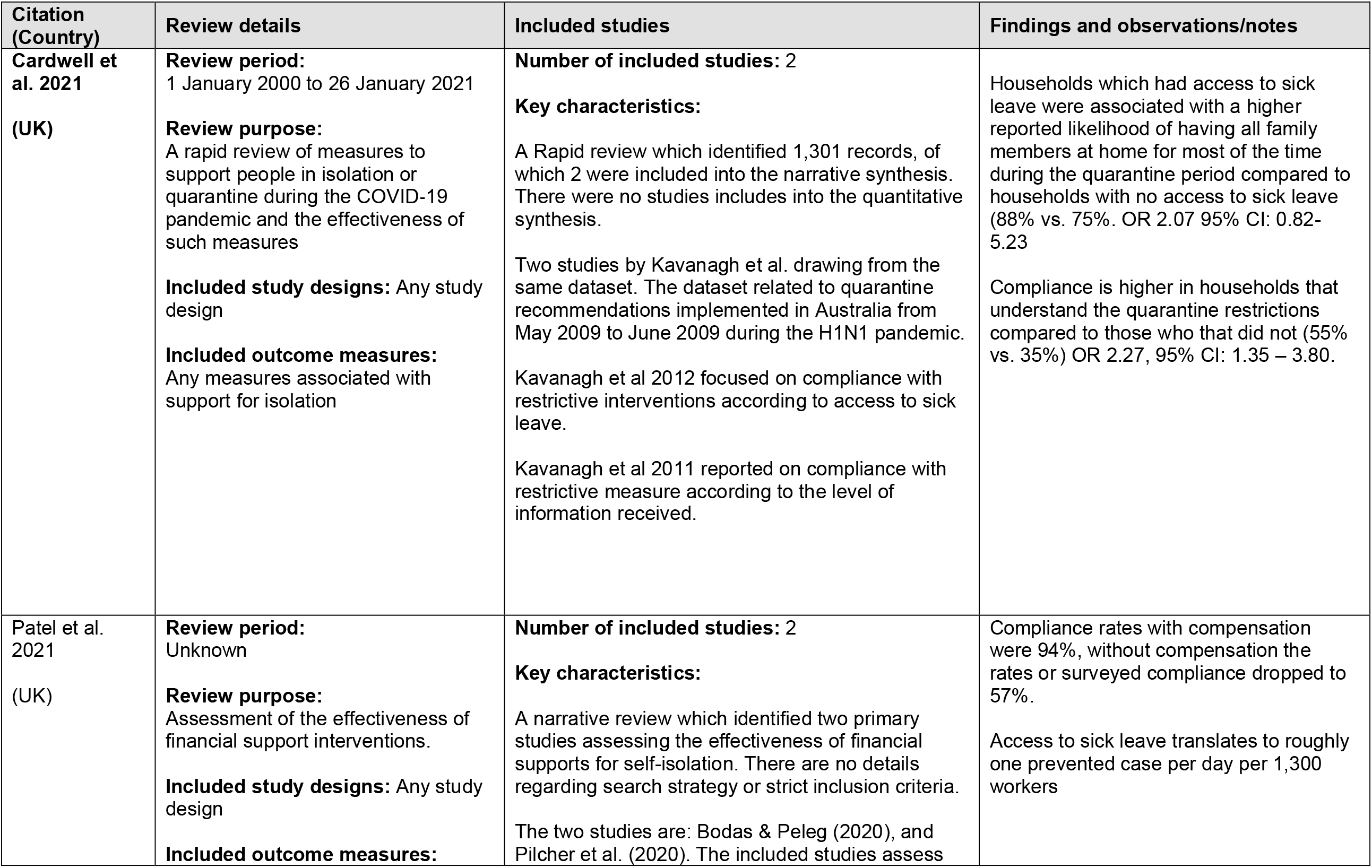
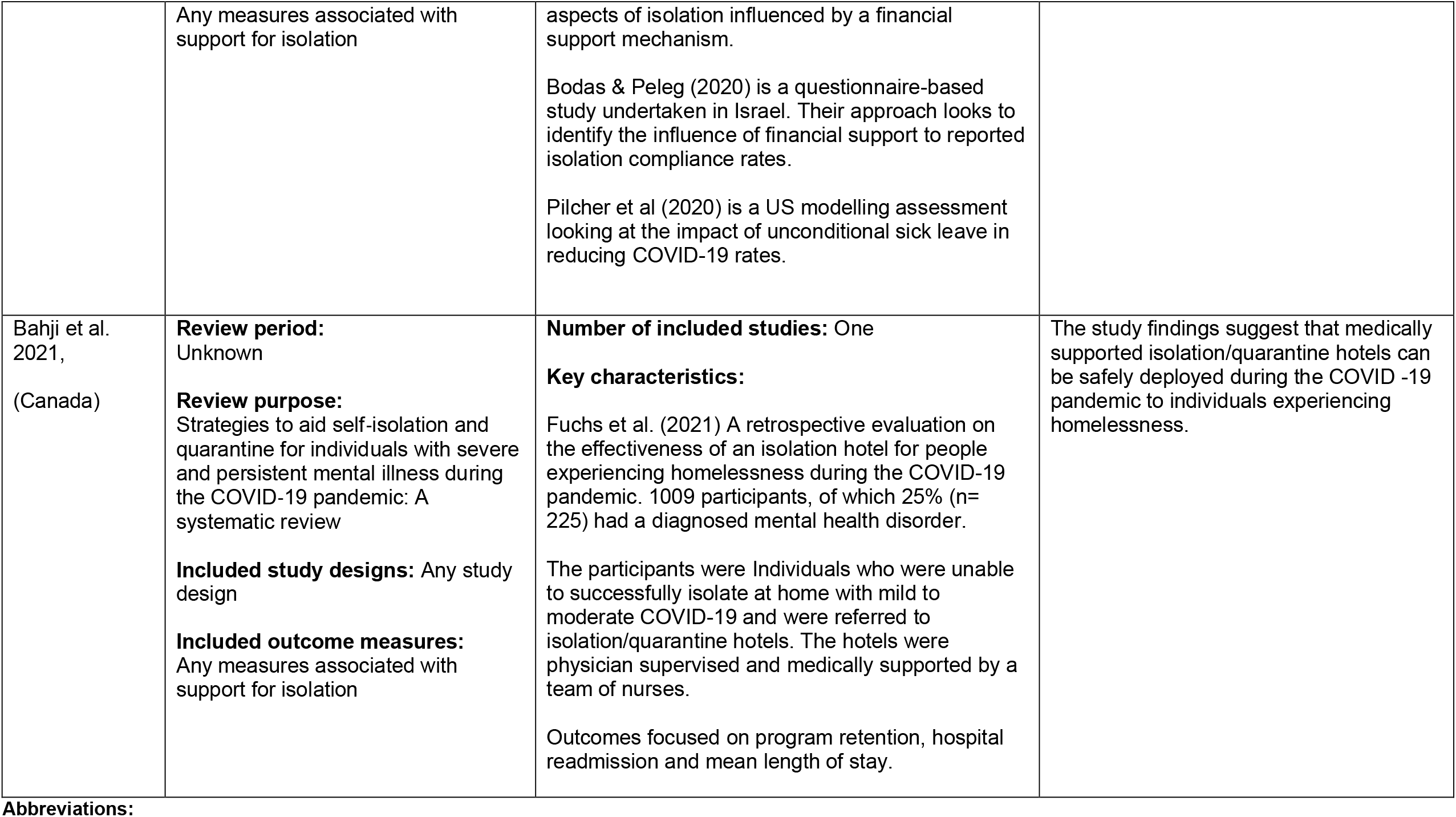
Summary of rapid reviews.

**Table 2:** Summary of primary studies.

| Citation (Country) | Study Details | Participants & setting | Key findings | Observations/notes |
| --- | --- | --- | --- | --- |
| **Primary studies identified from reviews** |  |  |  |  |
| **Bodas & Peleg (2020)(Israel )** | **Study design:** Questionnaire<br>**Type of intervention:** Assessing public preference to aspects of COVID-19.<br>**Data collection Methods:** Online polling service (iPanel). | **Sample size:** 563<br>**Participants:**<br>Cross sectional questionnaire in Israel. Respondents mean age 39.6 years, with a minimum age of 18 years old. Male 50%, Jewish 80%. Employed 73%.<br>**Setting:** Israel, population level<br>**Dates of data collection:** Last week of February 2020 | **Primary findings:**<br>When offered financial support the level of isolation compliance was reported to be 94%, without financial support this was reduced to 57%.<br>The level of individuals reporting that they would not comply with isolation regulations (2 weeks) was 0.7% when monetary compensation was offered for lost wages, this increased to 11% when without support. The remaining percentage from each self-isolation circumstance was the option 'don't know or maybe'.<br>**Limitations:**<br>Whilst the study was conducted during the pandemic it is noted that situational attitudes may change as the pandemic progresses. The questionnaire was internet-based, the requirement of sufficient computer literacy may exclude a portion of the population and make the findings non-generalisable. The questionnaire asks respondents to predict future behaviour, this may not be as accurate as reporting based on action. The findings may not be generalisable beyond the Israel context. |  |
| **Kavanagh et al. (2012)(Australia)** | **Study Design:** Household survey data collection<br>**Type of intervention:** Access to sick leave and the associated | **Sample size:** 133 households<br>**Participants:** households with both parents employed. Participants identified through schools; each household has children of school age. | **Primary findings:**<br>Households which had access to sick leave were associated with a higher reported likelihood of having all family members at home for most of the time during the quarantine period compared to households |  |
|  | compliance to isolation during the H1N1 pandemic<br>**Data collection methods:** survey | **Setting:** School linked questionnaire<br>**Data collection dates:** November and December 2009 | with no access to sick leave (88% vs. 75%. OR 2.07 95% CI: 0.82-5.23<br>**Limitations:**<br>Lack of generalisability to the COVID 19 pandemic.<br>Uncertainty as to current behavioral preferences due to age of the research.<br>The findings may not be generalisable beyond the Australian context. |  |
| **Kavanagh et al. (2011)(Australia)** | **Study Design:** Household survey data collection<br>**Type of intervention:** Information dissemination to families who have entered home quarantine associated compliance to isolation during the H1N1 pandemic.<br>**Data collection methods:** survey | **Sample size:** 133 households<br>**Participants:** households with both parents employed. Participants identified through schools; each household has children of school age.<br>**Setting:** School linked questionnaire<br>**Data collection dates:** November and December 2009 | **Primary findings:**<br>Compliance is higher in households that understand the quarantine restrictions compared to those who that did not (55% vs. 35%) OR 2.27, 95% CI: 1.35 – 3.80.<br>**Limitations:**<br>Lack of generalisability to the COVID-19 pandemic.<br>Uncertainty as to current behavioral preferences due to age of the research.<br>The findings may not be generalisable beyond the Australian context. |  |
| **Fuchs et al. (2021)(USA)** | **Study design:** Retrospective observational study<br>**Type of intervention:** medically supported isolation/quarantine hotel for people experiencing homelessness.<br>**Data collection methods:**<br>Routinely collected data associated with the delivery of a novel isolation support intervention. | **Sample size:** 1,009 hotel guests<br>**Participants:** Individuals with the requirement for isolation who were unable to safely isolate at home. Mean age 44, 45% male. 346 transferred from hospital.<br>**Setting:** San Francisco, California.<br>**Data collection dates:** 19 March to 31 May, 2020 | **Primary findings:**<br>The study findings suggest that medically supported isolation/quarantine hotels can be safely deployed during the COVID -19 pandemic to individuals experiencing homelessness. Authors reported that 95% (776/955 guests) completed their stay, and 5% discontinued voluntarily (this analysis excluded guests who were discharged for other reasons, such as needing higher level of care or safety reasons.<br>**Limitations:**<br>Lack of generalisability to broader populations. |  |
|  |  |  | The findings may not be generalisable beyond the local American context. |  |
| **Pichler et al. (2020)(USA)** | **Study Design:** Epidemiological modelling study<br>**Type of intervention:**<br>Guaranteed universal sick pay, families first coronavirus response act (FFCRA)<br>**Data collection methods:**<br>Collated routine data sources | **Sample size:** 12 states enacted emergency sick pay (as well as district of Columbia) 38 states with no emergency sick pay mandate.<br>**Participants:** Population data including: Daily new infections, daily tests performed, policy measure enacted, state level population counts, state and city level sick pay mandates.<br>**Setting:** United states, state level isolation compliance and subsequent changes to infection levels.<br>**Dates of data collection:** 08 March 2020 to 11 May 2020. | **Primary findings:**<br>When accounting for a range of confounders, FFCRA was associated with a reduction of around 400 cases per day per state compared to states which didn't have FFCRA rules applying. This reduction amounts to a single case per day per 1,300 workers who received the option to take two weeks of paid sick leave.<br>**Limitations:** Analysis was undertaken using a short duration of data, this may not be representative of longer-term trends. Whilst a range of confounders were accounted for in the model, there may be unobserved factors influencing the changes in infection rates. There remains uncertainty as to the mechanism of effect for sick pay. |  |
| **Additional primary studies published after the reviews** |  |  |  |  |
| **Albani et al. (2021)(Brazil)** | **Study Design:** Epidemiological modelling study<br>**Type of intervention**<br>**[exposure]:** Relative level of 'Auxílio Emergencial' (AE) a social financial support program. The comparison is based on the availability of AE during the initial time of interest and the subsequent period where AE was not available (pre- and post-comparison).<br>**Data collection methods:**<br>Routinely collected data. | **Sample size:** National evaluation in Brazil. COVID-19 comparisons across states according with focus on AE and social isolation levels.<br>**Participants:** National routine data study.<br>**Setting:** Brazil – focus on state level comparisons.<br>**Dates of data collection:** 1 Mar 2020 to 11 May 2021 | **Primary Findings:** Financial support was associated with an increase in social isolation and a reduction in COVID-19 transmission and mortality. During the first wave, the AE was fully operational, this was a period where significantly higher social isolation was observed. The authors conclude that AE helped people adhere to social isolation rules and aided a reduction in the spread of the virus.<br>**Additional Findings:**<br>The authors estimated the incremental impact reductions in social isolation index | All statistical analysis is available on GitHub, datasets are well documented. |
|  |  | Two comparison periods. April to August 2020 (wave 1) and January to March 2021 (wave 2) | **Limitations:**<br>Confounding factors are not adjusted for in the analysis; for example, seasonality is not accounted for, there may be different risks associated with social contact relating to seasonal behaviours.<br>There may be generalisability issues from AE to other types of financial support for isolation.<br>There may be an issue of transportability of findings from the Brazil setting to other countries.<br>The two periods assessed within the comparison may be biased by the initial novel nature of the pandemic restricting social isolation followed by the period where there was a lack of for COVID-19 fear or psychological saturation to isolation. |  |
| **Bourdeaux et al. (2021)(USA)** | **Study Design: Economic modelling**<br>**Type of intervention**<br>[**exposure**]: Financial support for isolation<br>**Data collection methods:**<br>Routinely collected data. | **Sample size:** Massachusetts level routinely collected data.<br>**Participants:** State level (Massachusetts) modelled population.<br>**Setting:** Population aggregate level: Massachusetts (USA) potential isolation support finance program.<br>**Dates of data collection:** modelling period 01 February 2021 to 31 August 2021. | **Primary Findings:** The costs associated with deploying financial support for individuals required to self-isolation was estimated to be approximately \$430 per person. A reduction of the average reproduction rate from 1.09 to 1.08 was deemed to result in the programme being cost effective, when the medical cost average was \$2,500 per case.<br>**Limitations.**<br>There are multiple influential assumptions that are included in the economic models. The clinical effectiveness of financial support for self-isolation is included to illustrate a potential transmission reduction and subsequent cost impact, it is an assumption. The cost per person outcome has lower generalisability to other settings. | |
| **Johnson et al. (2022)(USA, Mexico, and Kenya.)** | **Study Design:** Discrete choice experiment (DCE)<br>**Type of intervention:** Preference ranking of non-pharmaceutical COVID-19 mitigation measures.<br>**Data collection methods:** Online DCE questionnaires. | **Sample size:** 71 participants<br>**Participants:** University employees or students from a range of World Bank income groups: High income (USA) upper middle (Mexico) and Lower missile income (Kenya). 18 years or older.<br>**Setting:** USA, Mexico, and Kenya<br>**Dates of data collection:** Questionnaire September – November 2020 recalling preferences from the initial wave of COVID-19, April – June 2020. | **Primary findings**<br>Findings suggest that participants preferred to shelter in place for a greater number of days a week to have a lower COVID-19 risk. There was a preference to receive financial government compensation during the pandemic. Both findings were statistically significant.<br>**Additional information:**<br>DCE design – nine individual-level attributes based on experiences during the initial three months of the COVID-19 pandemic (April 2020 to June 2020). The range of attributes included both subjective and objective components.<br>**Limitations:**<br>Low sample size. Insufficient sample size to disaggregate by country. Potential for recall bias due to duration from period of interest. |  |
| **Homaie Rad et al. (2021) (Iran)** | **Study Design:** Discrete choice experiment (DCE)<br>**Type of intervention:** DCE looking at the willingness to accept being isolated for COVID-19 in Iran.<br>**Data collection methods:** Online DCE questionnaires sourced by social network. Fourteen choice sets with two scenarios in each. | **Sample size:** 617 participants<br>**Participants:** 60% female. 14% less than primary school degrees, 7% secondary school education, 39% high school education and 40% university degree education. Authors concluded that their sample was broadly representative of the Iranian population.<br>**Setting:** Iran<br>**Dates of data collection:** Questionnaire March to May 2020. | **Primary findings:**<br>People preferred higher payment for being isolated for a longer duration of days. The willingness to accept amount for seven days of isolation was US\$51.71. There is variation in financial preference across different socioeconomic, health and demographic status.<br>**Additional information:**<br>Financial incentives must cover all household members to have optimal COVID-19 prevention.<br>**Limitations:** | |
|  |  |  | Lack of geographic generalizability. |  |

We identified a further four studies published after the review searches were undertaken (Albani et al. 2021, Bourdeaux et al. 2021, Homaie Rad et al. 2021, Johnson et al. 2022). Alongside the five studies identified from reviews, a total of nine primary studies are included in this report. Most of the included studies focused on the COVID-19 pandemic (n=7), with two publications set in the context of the H1N1 pandemic. The study types included: epidemiological modelling studies (n=2), an economic modelling study (n=1), a questionnaire-based publication (n=1), discrete choice experiments (n=2), a retrospective observational study (n=1), and household surveys (both H1N1 studies, n=2). A summary of all included primary studies is provided in Table 2 and their findings summarised in Section 2.

We also identified one ongoing systematic review by Mendonca et al. (2020), which aims to identify barriers and facilitators to population level adherence to COVID-19 prevention measures. The review focuses on qualitative methods and synthesis. The inclusion criteria are adults who have received protective behaviour recommendations related to a pandemic. A summary of the review protocol is provided in Table 3. No results have yet been reported for this review.

**Table 3:** Summary of ongoing systematic reviews.

| Citation<br>(Country) | Study Details | Participants & setting | Key findings | Observations/notes |
| --- | --- | --- | --- | --- |
| **Mendonca et al. (2020)(USA)** | **Study design:** rapid qualitative evidence synthesis protocol<br>**Research aim:** to summarise and evaluate the evidence on barriers and facilitators to population adherence to prevention and control measures in COVID-19 and other respiratory infectious diseases<br>**Methods:** A systematic search of Medline, Embase and PsycINFO for studies on adults who are required to take protective action during a pandemic. | **Inclusion criteria:**<br>Adult population receiving protective behaviour recommendations. Qualitative data collection **and analysis methods or mixed methods approaches.**<br>Studies published at any time in English, Portuguese, and Spanish<br>**Exclusion criteria:**<br>Studies not analysing data with qualitative methods; systematic reviews, books, policy reports, editorials, letters to the editor, conference papers, abstracts or expert reviews and grey literature. | No results published at time of this report.<br>**Strengths:**<br>Publication of protocol<br>Adherence to PRISMA process.<br>Two author sifting<br>Quality assessment<br>**Limitations:**<br>Language limits may restrict results<br>Patient and public involvement not involved in methods. |  |

**Table 4.** Eligibility criteria.

|  | Inclusion criteria | Exclusion criteria |
| --- | --- | --- |
| **Participants** | Anyone requested to self-isolate due to a positive COVID test or positive contact |  |
| **Settings** | Pandemic population level |  |
| **Intervention / exposure** | Any financial scheme developed with intention to support self-isolation |  |
| **Comparison** | No Support or standard level of support |  |
| **Outcomes** | Impact on virus transmission<br>Impact on compliance with self-isolation<br>Impact on inequalities |  |
| **Study design** | Any study design |  |
| **Countries** | Any country |  |
| **Language of publication** | English language |  |
| **Publication date** | Post 1<sup>st</sup> Jan 2021 for main search – structure builds on two recent rapid reviews |  |
| **Publication type** | Published and preprint |  |

### 2.2 Effectiveness of financial support for isolation

#### 2.2.1 Evidence from epidemiological modelling studies

Two epidemiological modelling studies evaluated whether there was a reduction in COVID-19 transmission due to an increase in social isolation (Albani et al. 2021, Pichler et al. 2020). Pichler et al. (2020) estimated the impact of unconditional sick leave to support individuals who may need to self-isolate in the USA. Their state level analysis was used to compare outcomes for states where the intervention of sick leave associated with the families first coronavirus response act (FFCRA) had been enacted with states that had not implemented the act during the assessment period of March to May 2020. The model included data on new daily infections, daily testing, sick leave policies enacted, and state population size. A reduction in daily new infections was estimated to be associated with the FFCRA availability of sick leave. The scale of the effect is estimated as 400 fewer cases per day for states implementing FFCRA. The reduction in new infections meant that for every 1,300 workers who received unconditional sick pay, there was one fewer new infection per day. The authors noted that a large number of potential confounders were not accounted for by the model.

Albani et al. (2021) used mobile phone data in order to identify the effectiveness of financial supports (Auxílio Emergencial [AE]) in reducing COVID-19 transmission in Brazil. State level social financial support data was paired with social isolation scores to identify a relationship between financial support and higher levels of social isolation. A pre- and post-social financial support comparison was adopted. It estimated that the higher level of social isolation seen during the period that AE was available was associated with a reduction in viral spread. The authors highlight the limitations of their study and potential confounding factors that may influence the results, such as potential behavioural changes in different phases of the pandemic and due to the initial compliance being motivated by the emergence of a deadly disease. The later period may be characterised with lower social isolation due to the psychological saturation of having endured prior periods of the pandemic. Changes in season may also be a confounding factor.

#### 2.2.2 Evidence from economic modelling studies

Bourdeaux et al. (2021) developed an economic model to compare the cost of delivering financial support and the cost of inaction. Their model setting was the Massachusetts state population with employment characteristics and COVID-19 transmission figures reflecting this population. The modelling period was between 1 February 2021 and 31 August 2021, and estimated costs to support a quarantine programme to be $430 per person. The total estimated cost of the financial support was estimated to be between $300m and $570m and would cover the 800,000 to 1.3 million Massachusetts residents through to August 2021. Their cost-effectiveness equation assumed that the increase in social isolation may result in a reduction of 0.01 to the reproduction rate from the figure of 1.09 that was measured at the time of developing the model. There was no rationale provided to support the 0.01 figure. Under the assumption that social isolation would reduce the reproductive rate by 0.01, the authors found the intervention offer a possible reduction in cases of 100,000, with 1,800 fewer deaths and a reduction in direct medical costs of $265 million.

#### 2.2.3 Evidence from questionnaire-based publications

Bodas & Peleg (2020) reported findings from an internet-based questionnaire on attitudes towards various COVID-19 related factors. The Israel-based survey included questions on the likelihood of isolation compliance (2 weeks) according to whether government financial support to compensate for lost wages was available or not. Findings suggested that compliance with isolation increased if financial support was offered, with 94% of individuals reporting an intention to comply when offered finance compared to 57% when compensation was not available.

#### 2.2.4 Evidence from Discrete choice experiments

Johnson et al. (2022) and Homaie Rad et al. (2021) undertook discrete choice experiments (DCEs) to elicit public preferences regarding COVID-19 in the United States of America and Iran, respectively. Johnson et al. (2022) undertook their DCE across universities in the USA, Mexico and Kenya to try to represent an income spread in their findings. Respondents reported a preference to receive financial government compensation during the pandemic. In addition to preferring financial support, individuals were willing to shelter in the same place for a longer duration in order to reduce their COVID-19 risk. Homaie Rad et al. (2021) constructed a willingness to accept framework within the DCE in order to identify levels of financial support where individuals were indifferent when accepting a 7-day isolation. The amount of US dollars individuals would require in order to accept a 7-day isolation was $51.71.

#### 2.2.5 Evidence from retrospective observational studies

Fuchs et al. (2021) undertook a retrospective cohort study looking at the impact offered by a medically supported isolation hotel for people experiencing homelessness in the USA. They reported outcomes from 1,009 hotel guests who were unable to safely isolate at home during the period 19 March to 31 May 2020. The authors included 955 of the 1,009 guests in an analysis of retention and voluntary premature discontinuation; 54 guests were excluded from the analysis due to other discharge reasons. Overall, 95% of guests completed their quarantine stay (776 of 955 guests). These findings suggest that medically supported isolation/quarantine hotels can be safely deployed during the COVID-19 pandemic to individuals experiencing homelessness.

#### 2.2.6 Evidence from the H1N1 pandemic

Kavanagh et al. published two papers, one in 2011 and a second in 2012, both focused on the H1N1 pandemic. The papers report findings from an Australian household survey questionnaire. Kavanagh et al. (2011) found that households that received financial support reported a higher likelihood of having all of their family members at home for most of the quarantine period compared to the situation where no financial support was offered (88% versus 75%, respectively). Kavanagh et al. (2012) identified an increase in compliance likelihood according to improved information regarding understanding of restrictions.

### 2.3 Bottom line results for effectiveness of support for isolation

A limited evidence base suggests that financial support for isolation is associated with an increase in isolation compliance, lower social engagement, and a reduction in infection levels. The hypothesised causal pathway between the implementation of financial support for isolation and a reduction in COVID-19 cases is well documented, however, the extent to which the components interact is uncertain. There is evidence suggesting individuals are more likely to comply with isolation when they are financially supported to do so (Albani et al. 2021, Bodas & Peleg 2020, Homaie Rad et al. 2021, Johnson et al. 2022, Kavanagh et al. 2012, Pichler et al. 2020). Evidence suggesting a positive relationship with longer isolation duration and higher financial requirements (Homaie Rad et al. 2021). Increased levels of social isolation are associated with reduced transmission of COVID-19 in epidemiological models (Albani et al. 2021, Pichler et al. 2020). Financial support programmes have the potential to be a cost-effective intervention (Bourdeaux et al. 2021). In addition to the evidence on epidemiological and cost-effectiveness, Fuchs et al. (2021) offered evidence supporting the viability of delivering medically assisted isolation hotels for people unable to isolate at home. Further to the COVID-19 literature, two studies found that a financial support and improved social restriction information was associated with compliance with H1N1 isolation (Kavanagh et al. 2011, Kavanagh et al. 2012). However, direct real-world evidence on the implementation of financial support for COVID-19 and its effectiveness on self-isolation compliance and subsequent reduction in SARS-CoV-2 transmission, is limited.

## 3. DISCUSSION

### 3.1 Summary of the findings

This rapid evidence review aimed to identify evidence on the effectiveness of financial support for people who are required to isolate. Multiple questionnaire-based publications have reported higher isolation compliance because of financial support. Two epidemiological modelling studies have observed higher levels of social isolation in locations which made finance for isolation/sick leave accessible compared to those without support. A major limitation with this literature is that the studies either rely on reported behaviours or, where observed levels are assessed, there is a range of confounders not accounted for. Whilst there are multiple publications concluding that financial support is associated with a higher compliance to isolate, there remains considerable uncertainty as to its effectiveness on transmission.

There is limited evidence to suggest that financial support improves isolation compliance and reduces COVID-19 transmission levels. The two epidemiological studies, which observed higher social isolation, both suffer from short data collection periods, uncontrolled confounders, generalisability concerns and issues with seasonality. As with support for isolation compliance, there is uncertainty as to the extent to which financial support impacts transmission.

The nature of assessing the effectiveness of an intervention during a dynamic pandemic has meant that the certainty with which we can conclude effectiveness is relatively low. For studies that utilise observed trends and outcomes there is a consistent lack of appropriate comparator and the risk of unobserved confounders. Studies which undertook a questionnaire approach must accept the uncertainty related to behavioural adaptation to the constantly moving pandemic setting.

There was no evidence related to effectiveness of financial support for disadvantaged populations who are required to isolate or any insight to the impact of financial support on equality.

### 3.2 Limitations of the available evidence

The evidence base consists of a small collection of studies with no randomised control trials (RCTs). The range of limitations inherent in the current literature can be broadly broken up into structural study limitations and poor generalisability of findings. The structural limitation heading includes the factors associated with the undertaking of the research, such as, inappropriate comparator, unobserved confounders, and hypothetical scenarios.

The pandemic setting offers an ever-changing landscape for researchers to try to isolate causal inference. The lack of well controlled studies means that the study designs risk having inherent bias. Whilst there were efforts taken to offer appropriate comparators within the comparative studies, it is difficult to control for potential differences across timelines or geographies. Findings were adjusted according to the relative characteristics of the comparator, however, there, was a high likelihood of unobserved confounding. Confounding effects which are not accounted for may influence the outcomes of interest and introduce bias.

Most primary studies in this review are based on questionnaire response assessments. The central issue with questionnaire studies is that the outcomes are reported as opposed to observed. Participants may signal a proposed behaviour which doesn’t correlate with observed action. In addition to this central limitation, questionnaire responses may suffer from responder agenda, in this case, wanting to support a policy to offer financial support.

The second limitation area is that of generalisability. There is uncertainty as to the likelihood that the conclusions drawn from the literature would be observed in another time period, location or population. The pandemic is dynamic, whether that is current infection levels, dominant COVID-19 variant, season, vaccination coverage or non-pharmaceutical intervention status. Findings based on data collected on a sub sample of the pandemic duration may find that their conclusions are not observed when the study is repeated at a later date.

Location generalisability is a major limitation when drawing conclusions from a geographically disparate literature like the one included in this review. The variability of financial and social behaviours, social policy, and earnings is highly heterogeneous across the country settings for the studies. When assessing the impact of financial support, the country’s financial situation, distribution of finance within the country, behaviours towards finance, and current social support measures are important considerations. There may be population generalisability limitations in the included studies. The sample sizes reported are relatively small and may not reflect the true population preference.

### 3.3 Implications for policy and practice

Findings corroborate the use of financial support for isolation to increase isolation compliance, however there are caveats. Whilst the general finding that financial support is associated with a reduction in COVID-19 transmission there are significant limitations with the evidence base. There is no generalisable evidence to inform the optimal scale of financial support. Current evidence is insufficient to offer an insight into the scale of effectiveness of financial support for isolation may achieve. The most informative current evidence makes use of large routine data collections, it is possible that further research will offer more definitive and instructive conclusions.

### 3.4 Strengths and limitations of this Rapid Review

The studies included in this rapid review were identified using a systematic literature review of a range of carefully selected publication databases. The research question and study protocol were developed with significant input from experts in the field. The abstract and full text screening was conducted by a single researcher, uncertainty was checked by a second reviewer. The data extraction was performed by a single reviewer and checked for consistency by a second researcher.

Whilst the review methods undertaken have been pragmatically robust, there remains the possibility that additional eligible texts have been missed. The search was limited to English language publications which may have induced bias. We developed our search strategy and implemented a date restriction informed by the two rapid reviews that established the basis of this review. There may be eligible literature that was not identified in either review or was published prior to our date restriction.

The literature includes studies with serious limitations, whilst much of this is due to the difficulty of hypothesis testing within the COVID-19 context, this reduces the strength of conclusions. Efforts have been made to summarise the limitations of each study; however, this review undertakes no formal risk of bias assessments, therefore we are unable to say with appropriate context the strength of conclusions.

## Data Availability

All data produced in the present study are available upon reasonable request to the authors

## Abbreviations

Acronym: Full Description
COVID-19: Coronavirus disease 2019
CI: Confidence interval
OR: Odds ratio
DCE: Discrete choice experiment
FFCRA: Families first coronavirus response act
AE: Aux’
silio Emergencial (Financial support program)
RCT: Randomised control trials

## 5. RAPID REVIEW METHODS

### 5.1 Eligibility criteria

The focus of the report is on evidence relevant to the effectiveness of financial support for isolation during the COVID 19 pandemic, other pre-covid pandemics are considered.

### 5.2 Literature search

A systematic literature search was conducted between the 25^th^ – 29^th^ April 2022 across a range of databases for English language publications, and then 5^th^−6^th^ May 2022 for ongoing trials and ongoing reviews. The searches were restricted to studies published after 1^st^ Jan 2021 as the rapid reviews by Cardwell et al. (2022) and Patel et al. (2021) undertook searches of the prior literature. The search databases and search dates are listed in table 5. Appendix 1 documents the search strategy used for MEDLINE. Search strategies for other databases are available on request.

**Table 5.** Search databases.

| Database | Date Searched |
| --- | --- |
| Ovid MEDLINE | 26th April 2022 |
| Embase | 26th April 2022 |
| WHO Global Coronavirus database | 28th April 2022 |
| L*VE COVID | 28th April 2022 |
| Cochrane C-19 Study Register | 28th April 2022 |
| VA-ESP | 28th April 2022 |
| Cochrane Library | 28th April 2022 |
| Epistemonikos | 28th April 2022 |
| Scopus | 29th April 2022 |
| WHO ICTRP | 5th May 2022 |
| clinicaltrials.gov | 5th May 2022 |
| PROSPERO | 6th May 2022 |
| ABI Inform | 11th May 2022 |

### 5.3 Study selection process

The rapid reviews by Cardwell et al. (2022) and Patel et al. (2021) were identified during initial scoping searches, and form the base of the literature included in this review. As these rapid reviews align closely to the research question of this report, we searched for evidence published following the search terms from Cardwell et al. (2022) and Patel et al. (2021) (i.e. 2021 onwards). A further rapid review was identified, across the three rapid reviews there was a total of five primary studies with none appearing more than once in the reviews. Due to the scarcity of evidence, each primary study included in the three rapid reviews was extracted individually. Cardwell et al. (2022) included two studies reporting evidence from the H1N1 pandemic, aside form these two publications all evidence reports on the COVID 19 pandemic.

### 5.4 Data extraction

A single researcher performed the data extraction with consistency checks being carried out by a second researcher. The following information was extracted form secondary evidence:

- Citation
- Country
- Review period
- Review purpose
- Included study designs and outcomes
- Number of included studies
- Key characteristics
- Findings and observations

Data extraction from primary evidence was undertaken by the same methods as for secondary reviews, the information extracted was:

- Citation
- Country
- Study design
- Type of intervention
- Data extraction methods
- Sample size
- Participants
- Setting
- Dates of data collection
- Key findings

### 5.5 Quality appraisal

No formal quality appraisal was undertaken.

## 6. EVIDENCE

### 6.1 Study selection flow chart

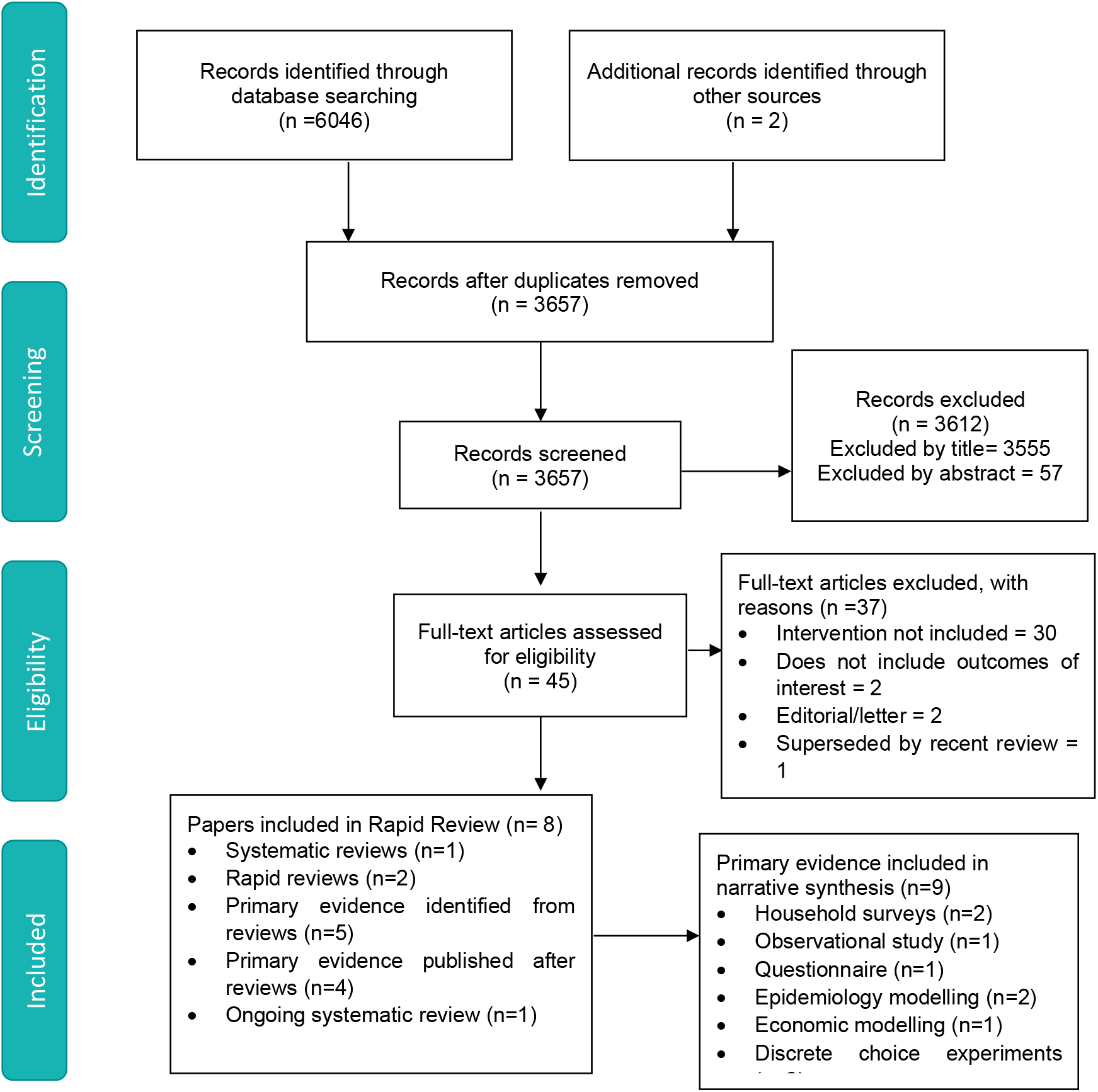

## 7.2 Acknowledgements

The authors would like to thank Nick Srdic, Jonathan Price, Sue Leake, Russell Williams and Chris Pavlakis for their contribution in guiding the focus of the review and to interpreting the findings.

## 8. ABOUT THE WALES COVID-19 EVIDENCE CENTRE (WCEC)

The WCEC integrates with worldwide efforts to synthesise and mobilise knowledge from research.

We operate with a core team as part of <u>Health and Care Research Wales</u>, are hosted in the <u>Wales Centre for Primary and Emergency Care Research (PRIME),</u> and are led by <u>Professor Adrian Edwards of Cardiff University.</u>

The core team of the centre works closely with collaborating partners in <u>Health Technology Wales</u>, <u>Wales Centre for Evidence-Based Care</u>, <u>Specialist Unit for Review Evidence centre</u>, <u>SAIL Databank</u>, <u>Bangor Institute for Health & Medical Research/ Health and Care Economics Cymru,</u> and the <u>Public Health Wales Observatory.</u>

Together we aim to provide around 50 reviews per year, answering the priority questions for policy and practice in Wales as we meet the demands of the pandemic and its impacts.

### Director

Professor Adrian Edwards

### Website

https://healthandcareresearchwales.org/about-research-community/wales-covid-19-evidence-centre

### 9. APPENDIX 1: MEDLINE search strategy

Database(s): **Ovid MEDLINE(R) ALL** 1946 to April 25, 2022

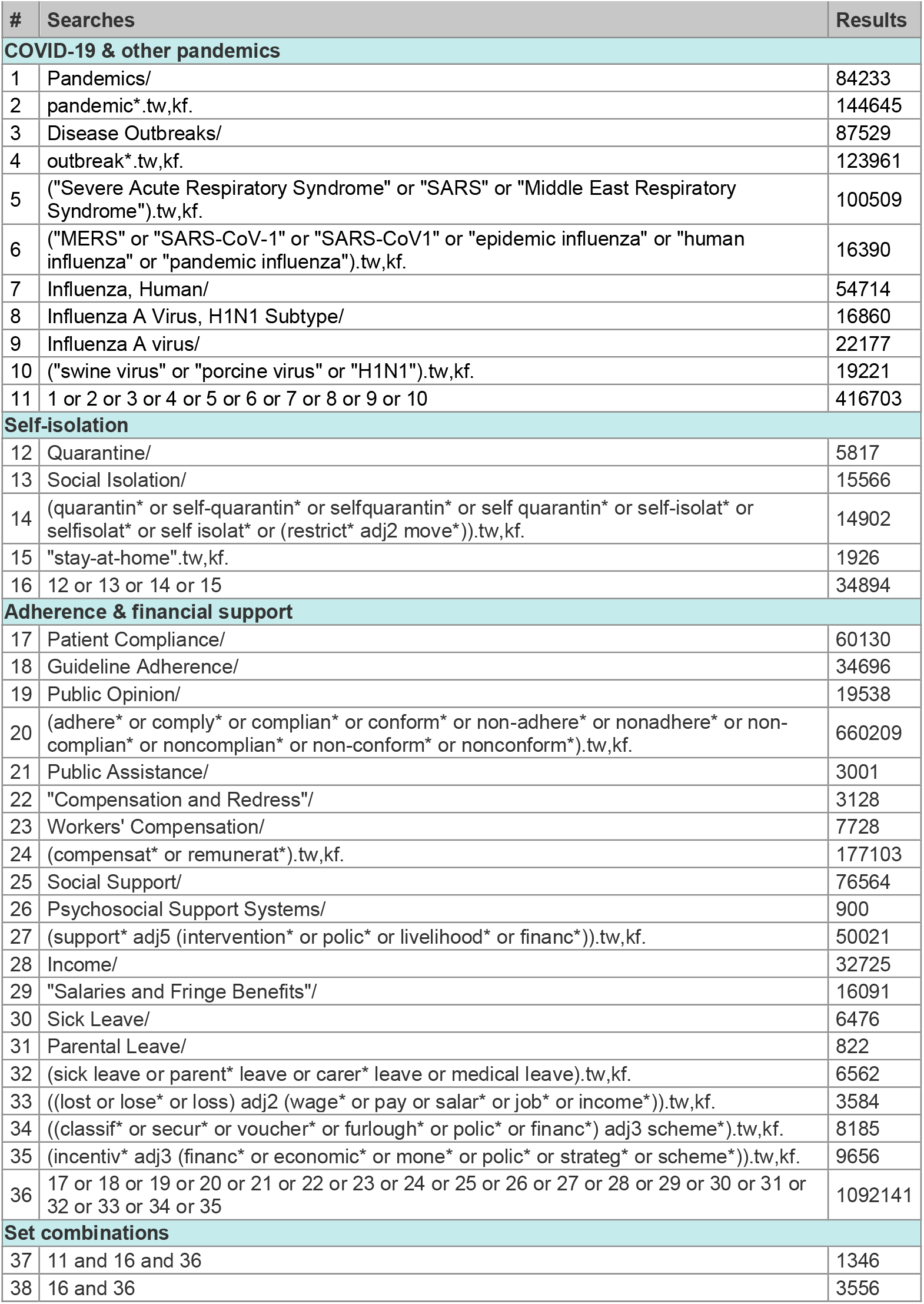
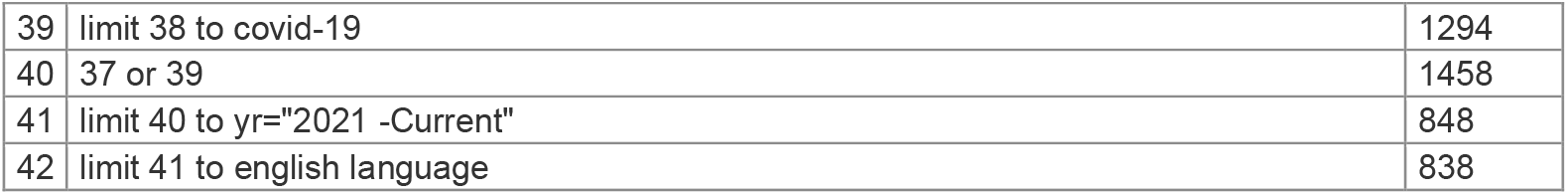

